# Sex-specific genetic regulation of proteomics in cerebrospinal fluid uncovers genetic causes for sex differences in neurodegeneration

**DOI:** 10.1101/2025.10.29.25339064

**Authors:** Soomin Song, Anh N. Do, Lihua Wang, Gyujin Heo, Jiseon Kwon, Daniel Western, Seung Jin Yang, Jigyasha Timsina, Menghan Liu, John Budde, Michael E. Belloy, Eric McDade, Merce Boada, Adelina Orellana, Maria Victoria Fernandez, Agustin Ruiz, Pau Pastor, John C. Morris, David Holtzman, Suzanne E. Schindler, Han Chen, Carlos Cruchaga, Yun Ju Sung

**Author notes:** Corresponding Author: Yun Ju Sung, PhD, Washington University School of Medicine, 4444 Forest Park Ave, Room 5501, Box 8134, St. Louis, MO 63108.

## Abstract

Sex-specific genetic regulation of cerebrospinal fluid (CSF) protein levels may contribute to differential vulnerability to neurodegenerative diseases. To systematically identify sex differences in the genetic regulation of CSF proteome and their link to neurodegeneration, we performed sex-stratified pQTL analysis of 6,361 proteins in 1,713 males and 1,640 females, separately. We identified 1,729 pQTLs significant in either sex. They included 407 sex-specific pQTLs (genetic regulation in only one sex) and 159 sex-biased pQTLs (regulation in both sexes, but with different magnitudes of regulation between sexes). The *HLA* locus on chromosome 6 and the *APOE* locus on chromosome 19, two known pleiotropic regions, regulated several proteins in a sex-dependent way. Pathway enrichment revealed several biological processes that were shared and distinctive of sex. Using proteome-wide association study (PWAS) and colocalization, we identified 22 proteins associated and colocalized with AD risk loci. TMEM106B and ACE proteins were identified in only one sex. Four proteins were associated and colocalized with PD risk loci. These findings provide insights into dissecting the underlying mechanisms contributing to sex differences in neurodegeneration.

## Introduction

Sex differences exist in the prevalence, onset, severity, and clinical features of multiple diseases including neurodegeneration. In Alzheimer’s disease (AD), the most common form of dementia, females have a greater life-time risk of developing AD than males^1^. After diagnosis, females experience faster disease progression and more rapid cognitive and functional decline compared to males^2^. In contrast, Parkinson’s disease (PD) shows a male predominance, with incidence and prevalence approximately 1.5 times higher in males^3^. Males also experience symptom onset 2.2 years earlier. Proposed hypotheses underlying these discrepancies include differences in life expectancy, sex-related hormones, and sex-specific immune reactions^4,5^. However, the underlying molecular mechanisms driving sex-specific disease vulnerability are unclear.

To bridge these gaps between sex differences in disease risk and underlying biology, many studies have examined sex differences in genetic regulation of disease risk and intermediate molecular traits, including transcriptomes and proteomes. Through genome-wide association studies (GWAS) with AD, female-specific associations with amyloidosis have been reported in genetic variants in serpin family members^6^. Male-specific associations with AD were reported in variants in *GRN,* a regulator of lysosomal function, and *TREM2,* which is critically involved in phagocytosis^7–9^. Through an expression quantitative trait locus (eQTL) study, the genotype-tissue expression (GTEx) project examined transcriptomic data across 44 tissues, identifying several sex-biased eQTLs in breast, muscle and adipose tissues^10^. Several of these eQTLs were colocalized with complex traits, explaining some of their sex differences. To examine genetic regulation on proteins, which are actively involved in biological processes, Wingo et al identified sex-specific protein QTLs (pQTLs) in the brain, linking them to psychiatric, neurological, and brain morphologic traits^11^. Koprulu et al examined sex differences in genetic regulation of the plasma proteome, reporting similar genetic architecture between sexes^12^. However, the relevance of the plasma protein involvement in brain-relevant traits is unclear due to the blood-brain barrier.

Cerebrospinal fluid (CSF), which surrounds the brain and spinal cord, can be collected from living individuals, offering a unique opportunity to study molecular mechanisms relevant to neurological diseases. We hypothesized that there would exist sex differences in the genetic regulation of CSF proteome, which may in turn help explain some of the sex differences observed in neurodegeneration. To test this hypothesis, we performed sex-stratified GWAS of 6,361 proteins in 1,713 males and 1,640 females, separately. We characterized the sex differences in pQTLs and identified the related biological pathways. We subsequently performed proteome-wide association studies (PWAS) and colocalization to identify proteins for AD and PD.

## Results

### Study overview and sample characteristics

To investigate sex differences in genetic regulation of the CSF proteome, we performed a sex-stratified GWAS of CSF proteome from 1,713 males and 1,640 females of European ancestry (Fig 1). Specifically, we considered the CSF proteome of these participants quantified by 7,008 aptamers (SomaScan 7K), which targeted 6,361 proteins. These aptamers were subsequently tested for associations with 12,630,196 autosomal variants that were imputed using the TOPMed reference panel. After obtaining significant pQTLs in either sex, we classified them into 1) sex-specific pQTLs, i.e., significant genetic regulations occurring only in one sex, and 2) sex-biased pQTLs, i.e., regulations occurring in both sexes but with different magnitudes of the effect (Fig 1; Supplementary Fig 1a).

This study included participants from six cohorts: the Knight Alzheimer Disease Research Center (Knight-ADRC), Alzheimer’s Disease Neuroimaging Initiative (ADNI), Fundació Ace (FACE), Bracelona-1, Dominantly Inherited Alzheimer Network (DIAN), and Parkinson’s Progression Markers Initiative (PPMI). The number of males and females across six cohorts was similar (∼51.1% of males; Table 1). They were middle-aged and older adults, with a mean age ranging from 38 to 74.5 years. Males and females exhibited no significant difference in age or APOE ε4 carrier status within each cohort.

**Table 1.** Demographic information of the samples included in this study.

| Cohort | Males |  |  | Females |  |  |
| --- | --- | --- | --- | --- | --- | --- |
| | N | Age | APOE ε4 N (%) | N | Age | APOE ε4 N (%) |
| Knight ADRC | 420 | 72.1 (8.61) | 161 (38.33) | 478 | 70.1 (8.54) | 190 (39.75) |
| ADNI | 418 | 74.5 (7.36) | 188 (44.98) | 315 | 72.3 (7.59) | 139 (44.13) |
| FACE | 183 | 72.4 (8.09) | 51 (27.87) | 267 | 71.3 (8.52) | 110 (41.2) |
| Barcelona-1 | 106 | 69.2 (6.91) | 42 (39.62) | 96 | 68.3 (8.21) | 38 (39.58) |
| DIAN | 111 | 39.4 (10.66) | 33 (29.73) | 136 | 38.3 (10.68) | 34 (25.37) |
| PPMI | 475 | 61.9 (10.28) | 117 (24.63) | 348 | 61 (9.35) | 68 (19.54) |

Knight ADRC = Charles F. and Joanne Knight Alzheimer Disease Research Center
ADNI = Alzheimer's Disease Neuroimaging Initiative
FACE = Fundació ACE Alzheimer Center
Barcelona-1 = Longitudinal observational study from the Memory and Disorder unit at Barcelona
DIAN = Dominantly Inherited Alzheimer Network
PPMI = Parkinson's Progression Markers Initiative
Age is age at Lumbar puncture (LP). Values denote years in mean (SD).
APOE ε4+ (%) denotes sample size of APOE ε4 allele carriers

### Sex differences in genetic regulations of CSF proteome

We identified 1,729 index pQTLs that were significant at least in one sex (1,502 pQTLs in males and 1,390 pQTLs in females; Fig 1; Supplementary Table 1). They consisted of 1,078 *cis*- pQTLs (62.3%; at P<5×10^-8^) and 651 *trans*-pQTLs (37.7%; P<3.5×10^-11^; Fig 2a). Among them, 1,163 pQTLs (67.2%) were significant in both sexes with no difference in their genetic regulation of protein levels between sexes (Supplementary Fig 1a). Specifically, the magnitudes of genetic regulations between males and females were highly correlated (r=0.99) at these shared pQTLs (Supplementary Fig 1b).

There were 566 pQTLs (32.7% of all index pQTLs) exhibiting some sex differences (Fig 2b-c). Among them, 407 were sex-specific (genetic regulation occurring in only one sex) and 159 were sex-biased (regulation in both sexes, but with different magnitudes of effect at P < 0.05). Of the pQTLs showing sex differences (either sex-specific or sex-biased), 52.9% were cis, a slightly lower proportion than in the overall pQTL results (62.3%), and they were more abundant in males (339 pQTLs, 59.9%) Among the sex-specific pQTLs, 230/407 were male-specific (cyan points in Fig 2b-c), with 109 *cis* and 121 *trans* associations. For instance, a *cis*-signal at rs12721109 (MAF=0.015) on chromosome 19 for SNRPD2 protein was significant in males (β =- 0.76, P=4.63×10^-8^) but not in females (β=-0.04, P=0.83; Fig 2d). Other well-known proteins including APOE, NEFL, and GRN had male-specific signals. Similarly, 177 pQTLs were female-specific (magenta points in Fig 1b). A *cis*-signal at rs71709684 (MAF=0.047) on chromosome 16 for IST1 was significant only in females (β=0.43, P=4.41×10^-8^ in females; β=-0.02, P = 0.79 in males; Fig 2d). Proteins including GBA, PPP3R1, and YWHAB had female-specific signals. Among the sex-biased pQTLs, 109 pQTLs were male-biased, exhibiting stronger genetic regulation in males. Proteins, such as FAM3B and CYB5D2, had male-biased signals. There were 50 female-biased pQTLs, including those for CEL and CLEC4G, exhibiting stronger genetic regulation in females.

The genomic annotations of identified pQTLs (Supplementary Table 2) were consistent among these shared, sex-specific, and sex-biased groups, predominantly present in intronic regions (25.2%-43%, Supplementary Fig 1c; Supplementary Table 3). The proportion of protein-altering variants (missense) ranged from 12.2% to 27.6% in each of groups. Consistent with published studies^13,14^, the magnitude of genetic regulations decreased as the locus moved farther from the transcription starting site (TSS) of their protein-coding genes (Supplementary Fig 2a). This pattern was similar between sexes (Supplementary Fig 2a). No significant differences were observed in the allele frequency of genetic variants associated with pQTLs between sexes (Supplementary Fig 2b). Finally, while most proteins (96%) were regulated by a single locus, a fraction of proteins were affected by multiple loci across genome. Among them, proteins including GNS and PTPA had both male- and female-specific pQTLs (Supplementary Fig 2c), suggesting complex sex-dependent genetic regulation.

### Validation and comparison with previous studies

To validate these identified genetic regulations of CSF proteome, we performed additional pQTL analysis, in 70 males and 95 females, separately, with CSF proteomic data (quantified with SomaScan 5K) from the Stanford ADRC cohort (Supplementary Table 4). Among 1,729 pQTLs identified in our primary analysis, 1,194 were available to examine in this Stanford proteomic data. Of these, 809 (67.8%) were nominally significant (P<0.05) in at least one sex. A high degree of directional concordance was observed between our primary analysis and these results, with an 85.2% agreement in males and 85.9% in females. Additionally, correlation of genetic effects between two results was also strong (r=0.85 in males, r=0.90 in females; Supplementary Fig 3), supporting validity of the identified pQTL results in CSF.

Given the biological relevance of CSF to brain, we further compared our results using previously published datasets. Specifically, we compared our findings to those from Wingo et al.^11^ which performed sex-stratified pQTL with mass-spectrometry proteomic data in dorsolateral prefrontal cortex from 263 males and 453 females (Supplementary Table 5). We observed high directional concordance (85.67% in males; 86.71% in females) even across brain and CSF. Correlations of genetics effects (r=0.63 in males, r=0.60 in females) were slightly reduced, potentially due to a different proteomic quantification and tissue heterogeneity (Supplementary Fig 4a).

Given a relevance of the proteome to transcriptomics, we next compared our results with two published sex-stratified eQTL results in brain, those from GTEx project^10^ and Wingo et al.^11^ The GTEx project^10^ performed sex-stratified eQTL analyses in 44 tissues (from 557 males and 281 females), out of which 13 brain regions were included. When we compared our results to these GTEx brain eQTLs (Supplementary Table 6), a moderate degree of directional concordance was observed with about 70% agreement (69.66% in males; 69.67% in females). Correlations of genetic effects were moderate (r=0.34 in males, r=0.38 in females; Supplementary Fig 4b). Next, we compared with brain eQTL results from Wingo et al.^11^ which performed eQTL analyses in dorsolateral prefrontal cortex samples from 213 males and 376 females. In this comparison (Supplementary Table 7), a bit higher directional concordance (74.37% in males, and 76.53% in females) was observed. Correlations of genetic effects were similar (r=0.36 in males, r=0.45 in females; Supplementary Fig 4c). While reduced correlation is likely due to differences in omic types (proteome vs. transcriptome) as well as tissue types (CSF vs. brain), high directional concordance supports some similarity between CSF pQTL and brain eQTLs.

### Sex differences in two pleiotropic regions

To identify genetic regulations shared across multiple proteins in CSF, we examined pleiotropic regions, genetic loci associated with multiple proteins in CSF, which may be important for brain function and disease. Out of 531genomic regions identified in this study, 132 were associated with at least three proteins (Supplementary Table 8). Among these, the *APOE* and *HLA* regions were the two most pleiotropic regions regulating many proteins. We subsequently performed a phenome-wide association study (PheWAS), pathway enrichment, and cell-type specificity analysis for the biological relevance of these two regions.

The pleotropic region on chromosome 6 is broad including 114 pQTLs (spanning from 24,503,369 to 33,809,483; chr6p21.1-p21.33; locusID=198 in Supplementary Table 1; Fig 3a), regulating 58 proteins in either sex (Supplementary Table 8). This is located around *HLA* (from 28,510,120 to 33,480,577). Proteins affected by this region were enriched with multiple biological processes involved in immune activities, including immune effector process and leukocyte and T-cell mediated immunity (Supplementary Table 9). Among the pQTLs identified in this region, 33 pQTLs showed sex differences. Specifically, we observed 18 genetic regulations only in males (male-specific) or more strongly in males (male-biased) for proteins including SCGN, SIRT5, and immunoglobulin component such as IGHD. Similarly, 15 genetic regulations were observed only or more strongly in females for proteins including HLA-C along with multiple surface proteins involved in immune system (such as CD8A, CD33, CD84), and interleukin receptors (such as IL15RA, IL18RAP). PheWAS results revealed the association of these sex-specific/biased pQTLs with rheumatoid arthritis, pulmonary disease, and the regulation of innate and adaptive immunity (Supplementary Table 10; Fig 3b). These proteins showing sex difference in their genetic regulations were enriched in microglia/macrophages (fold enrichment = 2.11 P = 1.79×10^-2^; Supplementary Table 11; Fig 3c), consistent with a previous study^13^. Together, these results support the importance of this region in immune-specific neurodegenerative outcomes.

The pleotropic region on chromosome 19 included 260 pQTL (spanning 44,819,487 to 45,702,991; chr19q13.32; locusID=500 in Supplementary Table 1), regulating the largest number of proteins (256 proteins; Supplementary Table 8; Fig 3d). This region included *APOE* (from 44,905,796 to 44,909,393), the strongest genetic risk factors for AD. In particular, the two variants of *APOE* (rs429358 and rs7412) genetically regulated 142 and 28 proteins, respectively. This is consistent to prior publications^13,14^. Pathway analysis suggested that proteins regulated in this region were involved in Crohn’s disease, infection, and multiple system atrophy, along with neurodegeneration (i.e., dystonia, frontotemporal dementia (Supplementary Table 12). Among the 260 pQTLs identified in this region, 101 showed sex differences. Specifically, we identified 60 pQTLs only or more strongly in males for proteins including NEFL, MMP3, and FOXO1. Similarly, 41 pQTLs were identified only or more strongly in females for proteins including YWHAE and PPP3R1. PheWAS results revealed the association of these sex-specific/biased pQTLs with lipid metabolism and inflammation processes in addition to dementia (Supplementary Table 13; Fig 3e). Interestingly, although APOE is primarily expressed in astrocytes, proteins that were differently regulated between sexes in this region were enriched in neuronal cells (Fold enrichment = 1.8, P = 1.26×10^-3^; Supplementary Table 11; Fig 3f), suggesting potential cell-cell communication between neurons and astrocytes. Together, these results support the role of the APOE region in contributing to neurodegenerative and psychiatric conditions.

### Biological pathways

To identify whether biological processes of proteins regulated in males were shared with those in females or distinctive, we performed cell-type specificity, pathway enrichment, and disease enrichment on two protein sets: first, with the 301 proteins genetically regulated only in males (male-specific) or more strongly in males (male-biased); and second, with 220 proteins regulated only (female-specific) or more strongly (female-biased) in females. The proteins with male-specific/biased pQTLs were enriched in endothelial cells (fold change =1.5, P=0.014; Fig 4a). Those proteins with female-specific/biased pQTLs show enrichment for oligodendrocytes (fold change=1.54, P=0.048).

Pathway enrichment analysis revealed four biological categories: 1) extracellular matrix (ECM), 2) lysosomal function, 3) signal transduction, and 4) immune system (Supplementary Table 14). The enrichment for the first three categories was observed in both sexes (Fig 4b-c), whereas immune system pathways were found only for proteins in females (FDR < 0.05; Fig 4b). Specifically, immune-related pathways enriched for female proteins included regulation of T cell proliferation and leukocyte proliferation. They were driven by immune mediators such as IL-10, a cytokine central to the neuroimmune interface, and VCAM-1, a vascular adhesion molecule. ECM-related pathways enriched for both sexes included extracellular membrane organization and ECM disassembly. They included MMP2, a matrix metalloproteinase involved in ECM degradation^15,16^, and PRSS2, a trypsinogen implicated in neuroinflammation^17^, both relevant to tissue remodeling and blood-brain barrier function. Lysosomal pathways included the lysosome and vacuolar lumen. They included GBA, a well-known PD risk gene^18^, and CLN5, known to be associated with neuronal ceroid lipofuscinosis and AD^19^. Signal transduction pathways included signaling receptor regulator activity containing GRN, BMP4.

Disease enrichment highlighted distinct sex differences (Supplementary Table 14). In males, enrichment for hypertriglyceridemia was observed, for which BMP4 and GRN were involved (Fig 4b). In females, enrichment for neurodegenerative and immune-mediated conditions was observed: presenile dementia (FDR = 1.91×10^-2^), intracranial aneurysm (FDR=1.65×10^-2^), and pneumonitis (FDR=3.02×10^-2^; Fig 4c). They included female-specific regulation of proteins such as AGER (RAGE), VCAM-1, MMP2, and IL-10. In particular, AGER, a receptor for advanced glycation end products, has been implicated in ALS, multiple sclerosis, and AD via promotion of oxidative stress and inflammation^20^.

### Proteins associated with AD and PD

To identify genetic regulation that may explain sex differences in two neurodegenerative diseases, AD and PD, we utilized CSF pQTLs identified in this study. Within each disease (AD and PD, separately), we considered two sets of pQTLs: results in males and those in females separately. In each combination, we performed two integrative analyses: (1) a proteome-wide association study (PWAS)^21^ to identify those proteins associated with disease risk and (2) colocalization to test whether two associations (one signal driving protein level and another driving disease risk) are driven by a single causal variant^22^.

First, through PWAS with AD GWAS^23^, we identified 36 proteins associated with AD (significant at FDR < 0.05; Supplementary Tables 15 and 16). While 29 proteins (including APOE and TREM2) were significant in both sexes, 7 proteins were significant in only one sex (Fig 5a; Supplementary Fig 5). Specifically, 6 proteins (IBSP, C1RL, APOBEC2, PTPA, TMEM106B, and GSS) were associated with AD only in males. Except for IBSP, their protein levels were positively associated with AD risk. There was one protein (DBNDD2) significant only in females. Second, through colocalization, we identified 27 proteins with evidence of a shared genetic signal with AD risk (at posterior probability of hypothesis 4, PP.H4 > 0.5; Supplementary Table 17; Fig 5b). Among them, three proteins (TMEM106B, PTPA, and IBSP) were colocalized with AD risk in males only. One protein (ACE) was colocalized with AD risk in females only.

In AD, 22 proteins were significant in both PWAS and colocalization approaches (Fig 5c). Among them, 18 proteins were significant in both sexes for AD loci, including *CD33, GRN, IL34, PILRA*, and *TREM2* (local plots in Supplementary Fig 6), indicating their consistent proteomic contribution to AD between sexes. The remaining 4 proteins showed sex differences. Only in males, TMEM106B, PTPA, and IBSP were both associated and colocalized with AD risk (Fig 5d). In particular, TMEM106B was positively associated and colocalized with AD risk in males (PWAS FDR = 4.00×10^-5^; coloc PP.H4=0.93), while no such evidence was observed in females (PWAS FDR = 0.244; coloc PP.H4=0.0017). ACE was somewhat complex. ACE was associated in both sexes (PWAS), with somewhat stronger effects in females (PWAS Z=-7.95, FDR = 2.12×10^-13^ in males; Z=-9.53, FDR = 2.15×10^-19^ in females). However, it colocalized in females only (PP.H4 = 0.99) as indicated by the proximity between the index variant (rs4277405) for female ACE protein levels and the index variant for AD GWAS at this locus (rs4292), while no such evidence was found in males (PP.H4 = 0.13; Fig 5d).

For PD, we performed the same integrative analysis with PD GWAS^24^. First, through PWAS analysis, we identified 10 proteins significantly associated with PD risk (Supplementary Tables 18 and 19). There were 4 proteins (ADAM15, FAHD1, EFNA1, AIF1) in males only, 1 protein (GBA) in females only, and 5 proteins (including FGCR2B, GPNMB) in both sexes (Fig 6a; Supplementary Fig 7). Second, through colocalization, we identified 8 proteins with evidence of a shared genetic signal with PD risk (Supplementary Table 20; Fig 6b): two proteins (ADAM15, GRN) in males only; 1 protein (GBA) in females only; and 5 proteins (including FCGR2B, GPNMB) in both sexes. Taken together, we identified 4 proteins that were significant in both approaches (Fig 6c). While FCGR2B and GPNMB were significant in both sexes (Supplementary Fig 8), ADAM15 and GBA were significant only in one sex (Fig 6d). ADAM15 was associated (PWAS Z = −4.17, FDR = 3.0×10^-3^) and colocalized with PD (PP.H4 = 0.97) in males, while no such evidence was found in females (PWAS FDR = 0.63, PP.H4 = 0.0007) (Supplementary Table 20). In contrast, GBA was associated and colocalized in females only (PWAS FDR = 4.41×10^-27^, PP.H4 = 0.99 in females; PWAS FDR = 0.85, PP.H4 = 0.047 in males).

## Discussion

To uncover sex differences in the genetic regulation of the CSF proteome, we performed sex-stratified pQTL analysis (in 1,640 males and 1,713 females, separately) with over 6000 proteins (SomaScan7K). We identified 1,729 pQTLs significant in either sex. These results were highly consistent in an independent CSF proteomic data from Stanford ADRC. There were 566 pQTLs (32.7%) exhibiting some differences between sexes. Of these, 407 pQTLs were detected only in one sex (sex-specific) and 159 were detected in both sexes but with different magnitudes of effects (sex-biased). The *HLA* and *APOE* loci regulated several proteins in a sex-dependent way. Pathway enrichment revealed several biological processes that were shared and distinctive of sex. Through integrative approaches, we identified 22 proteins associated and colocalized with AD risk loci, out of which TMEM106B, PTPA, IBSP, and ACE were identified in only one sex. Four proteins were associated and colocalized with PD risk loci.

This study identified 1,729 pQTLs in either sex. There were 67% (1,174) pQTLs with highly consistent genetic regulations between sexes (r = 0.98). The remaining 32.7% (566) exhibited some sex differences. These proteins regulated in a sex-dependent way included several proteins relevant to neurodegeneration, such as SNRPD2, APOE, NEFL, and GRN (with male-specific genetic regulation) and IST1, PPP3R1, and YWHAB (with female-specific regulation). Notably, SNRPD2 has been previously reported as sex-differentially regulated in both brain and blood^25^. Additionally, IST1 has been shown to produce different transcript isoforms in males and females, suggesting underlying sex-dependent regulatory mechanisms^26^. These findings indicate potential sex-dependent genetic architectures implicated in neurodegeneration. Additionally, some proteins such as GNS and PTPA were genetically regulated by multiple loci, including one male- and another female-specific locus. These findings underscore the complexity of sex-dependent genetic regulation, likely caused by interactions among genetic, hormonal, or epigenetic factors^27,28^.

A few studies investigated sex differences in genetic regulations for transcriptomics and proteomics. When our results were compared with pQTL results in the brain by Wingo et al,^11^ we observed a moderate correlation of genetic effects on proteome (about 0.60 in both sexes) with high directional concordance (86%), suggesting decent overlap of genetic regulation between CSF and brain proteomes. The consistency was further decreased when comparing to brain transcriptomic data with correlation around 0.36 and directional concordance around 70%, in both sexes in two independent eQTL studies (GTEx project^10^ and Wingo et al^11^). These findings are expected as the proximity between tissues (CSF vs. brain) and omics layers (proteome vs. transcriptome) decreases, the consistency reduces accordingly, likely reflecting distinct genetic regulation across tissues and molecular layers. These findings emphasize the importance of tissue- and molecule-matched datasets for sex-specific genetic regulation.

This study observed several sex-dependent genetic regulations at *HLA* and *APOE* loci, two pleotropic regions reported for both CSF and plasma proteome^13,14,29^. The *HLA* locus is known to be associated with T-cell responses, complement activation, and inflammatory markers. Supporting this prior knowledge, we found that several proteins including HLA-C, a key antigen-presenting molecule, and several immune surface proteins such as CD84, CD33, and IL15RA, had sex-dependent genetic regulation at this locus. Moreover, *APOE* haplotypes including rs7412 and rs429358 have been known to increase risk for neurocognitive impairment^30^, increase CSF phosphorylated and total Tau^31,32^, and decrease CSF Aβ42^32^ differently between males and females. We observed that several proteins regulated in this *APOE* locus in a sex-dependent manner were relevant to neurodegeneration, such as YWHAE, involved in synaptic function^33,34^, and NEFL, a marker of axonal degeneration^35^. These findings suggest that disproportional risk of AD between sexes may be related to possible sex differences in proteomic regulation and their subsequent contribution to neurodegeneration.

This study identified several biological processes that were shared and distinctive of sex. While signal transduction pathways were shared in both sexes, we noted the involvement of male-dependent proteins BMP4 and GRN that were previously implicated in neurodevelopment^36–38^. Male-specific regulation of BMP4, important in triglyceride metabolism^39^, identified in this study, aligns with reported sex-specific effects in BMP/TGF-β signaling pathways^40^. GRN (progranulin) was reported as sex-specific in frontotemporal dementia, amyotrophic lateral sclerosis, AD, and PD^36,37^. Our male-specific disease enrichment for hypertriglyceridemia aligns with these results, suggesting sex-dependent mechanisms in metabolic and neurovascular dysfunction. The most notable pathways identified in females were immune-related and lysosome pathways. The involved proteins were immune mediators such as IL-10 and VCAM-1, previously reported for microglial activation and neuroinflammation^41^, as well as multiple sclerosis and AD^42–44^. Similarly, they included lysosomal proteins CLN5 and GBA, previously implicated in neuronal ceroid lipofuscinosis, AD, and PD^18,19^. Extracellular matrix (ECM) pathways also included female-specific proteins, such as MMP2, previously linked to blood-brain barrier (BBB) disruption and AD^15,16^, and PRSS2, implicated in neuroinflammation.^17^ Female-specific enrichment for Crohn’s disease and pneumonitis identified in this study aligns with previously reported association of *IL-10* promoter polymorphisms with ulcerative colitis susceptibility only in female patients^45^. These findings highlight distinctive female-specific biological processes that may lead to increased vulnerability to neuroinflammatory and immune-related neurodegenerative processes.

Our integrated approach combining these pQTLs and AD GWAS through PWAS and colocalization identified 22 proteins that were related to AD, out of which TMEM106B, PTPA, and IBSP were significant only in males and ACE in females. Specifically, TMEM106B was significantly associated and colocalized with AD risk in males. TMEM106B is a transmembrane protein involved in lysosomal transport and function in neurons^46^. Alterations in *TMEM106B* gene were shown to be involved with the formation and accumulation of lytic vacuoles, particularly in neurons, and were linked to neurodegenerative disease risk^47–51^. *TMEM106B* variant has been associated with AD risk with evidence of regulation of *TMEM106B* expression in the human brain^52^. Consistent with the previous report of TMEM106B for AD risk in male Chinese Han populations^53^, our male-specific association of TMEM106B protein with AD risk indicates differential contribution to AD, likely through lysosomal function. In contrast, ACE, Angiotensin-converting enzyme, demonstrated a significant association with AD in both sexes (but with stronger effects in females) and female-specific colocalization. Involved in blood pressure regulation and electrolyte balance, this gene was previously associated with AD risk.^54–56^ Along with prior case reports of missense mutations in its association with early-onset or rapid progression of AD in female patients^57,58^, our findings suggest ACE-related contribution to sex-specific vulnerability in AD.

This study also identified four CSF proteins – ADAM15, FCGR2B, GBA, and GPNMB – associated with PD risk. Among them, ADAM15, A disintegrin and metalloproteinase 15, showed male-specific association and colocalization with PD risk. The ECM plays a crucial role in brain homeostasis and neuronal function, and disruption of ECM components has been implicated in neurodegenerative processes, including α-synuclein aggregation in PD^59,60^. ADAMs are known to contribute to altered BBB integrity and neuroinflammation^61^. While no prior study has directly linked ADAM15 to PD in a sex-specific way, emerging evidence from human BBB models suggests that astrocyte-mediated vascular dysfunction may differ by sex^62^. In contrast, the GBA protein showed female-specific association and colocalization with PD risk. As one of the well-known PD risk^63^, mutations in *GBA* reduce glucocerebrosidase activity and leading to lysosomal dysfunction and accumulation of toxic proteins^64–66^. Along with a recent sex-stratified genetic study presenting sex-specific differences in *GBA* variant carriers and familial parkinsonism^67^, our findings indicated the female-specific GBA association with lower risk of PD, possibly through a protection against lysosomal dysfunction in females.

There are several limitations in this study. First, our analysis examined autosomal variants, excluding the X chromosome. Incorporating sex chromosomes will identify additional sex differences in genetic regulations of protein levels and subsequently for disease susceptibility^68,69^. Second, we had limited success in the relevance of these pQTL for understanding sex differences in AD and PD. It is not clear whether this is driven by the proteomic coverage or the sample sizes. Future follow-up would be beneficial. Third, this study mainly focused on European ancestry individuals. Analyses in diverse ancestral populations will increase the generalizability of these findings to individuals of other ancestries.

In summary, this study performed a large-scale sex-stratified pQTL (in 1,640 males and 1,713 females, separately) with over 6000 proteins in CSF, uncovering distinctive sex-specific genetic regulations of the CSF proteome. Our findings complement sex differences in previous eQTL and pQTL studies. Integration of a sex-stratified CSF pQTL with AD and PD risk loci led to the nomination of several proteins showing sex differences. The pQTLs identified in this study may be useful for other neuropsychiatric or neurodevelopmental disorders including depression, schizophrenia, or frontotemporal dementia.

## Materials and Methods

### Study populations

We considered participants with CSF proteomic and genetic data from six cohorts: the Charles F. and Joanne Knight Alzheimer Disease Research Center (Knight ADRC), the Alzheimer’s Disease Neuroimaging Initiative (ADNI), the Fundació Ace Alzheimer Center Barcelona (FACE), Barcelona-1, the Dominantly Inherited Alzheimer Network (DIAN), and the Parkinson’s Progression Markers Initiative (PPMI). In total, 898 samples from Knight ADRC, 733 samples from ADNI, 450 samples from FACE, 202 samples from Barcelona-1, 247 samples from DIAN, and 823 samples from PPMI. A summary of sex-stratified sample sizes is provided in Table 1.

### Cohort demographics

#### Knight ADRC

The Charles F. and Joanne Knight Alzheimer Disease Research Center (Knight ADRC) at Washington University in St. Louis, established in 1985, is one of 36 NIH-funded ADRCs dedicated to advancing Alzheimer’s disease (AD) research with the ultimate goal of prevention and treatment. Through the Memory and Aging Project (MAP), participants undergo longitudinal clinical and psychometric assessments, imaging (MRI, PET), and biospecimen collection (including CSF), harmonized via the Uniform Data Set. Over 6,600 individuals have participated in Knight ADRC studies to date^70^. Additional details are available at https://knightadrc.wustl.edu.

#### Alzheimer’s Disease Neuroimaging Initiative (ADNI)

ADNI was launched in 2003 as a public–private partnership led by Michael W. Weiner, MD, with the aim of developing clinical, imaging, genetic, and biochemical biomarkers for early detection and tracking of AD^71^. Participants are recruited from >50 sites across the United States and Canada and undergo standardized neuropsychological assessments, structural and functional imaging, and biospecimen collection. Additional details are available at http://www.adni-info.org.

#### Fundació Alzheimer Center Barcelona (FACE)

Fundació ACE (FACE), headquartered in Barcelona, Spain, is a private non-profit institution established in 1995 to improve diagnosis, treatment, and research in AD. The center has collected 20,000 blood and 1,831 cerebrospinal fluid samples, analyzed over 10,000 genetic samples^72^, diagnosed over 30,000 patients, and contributed to nearly 150 clinical trials. Diagnoses are established by a multidisciplinary team using established international criteria. Further details are available at https://www.fundacioace.com.

#### Barcelona-1

Barcelona-1 is a longitudinal observational study carried out at the Memory and Disorders Unit of the University Hospital Mutua de Terrassa, Barcelona, Spain. Approximately 300 individuals were recruited at baseline, including those with Alzheimer’s dementia, non-AD dementias, mild cognitive impairment (MCI), or subjective memory complaints (SMC). Lumbar puncture and amyloid PET were conducted in participants with MCI, early-onset dementia (<65 years), or atypical presentations.

#### Dominantly Inherited Alzheimer Network (DIAN)

The Dominantly Inherited Alzheimer Network (DIAN), coordinated by Washington University in St. Louis, is a multinational family-based longitudinal study of autosomal dominant Alzheimer’s disease (ADAD). Mutation carriers and non-carriers undergo standardized assessments, including neuropsychological testing, brain imaging, and biospecimen collection (blood and CSF), to detect early changes associated with ADAD. Additional details are available at https://dian.wustl.edu.

#### Parkinson’s Progression Markers Initiative (PPMI)

The Parkinson’s Progression Markers Initiative (PPMI)^73^ is an observational, international study launched in 2010 with support from the Michael J. Fox Foundation to identify clinical, imaging, genetic, and biospecimen markers of Parkinson’s disease (PD). PPMI has enrolled about 4,000 deeply phenotyped participants with PD, prodromal PD, or healthy controls, and over 50,000 individuals for genetic screening. Additional details are available at www.ppmi-info.org.

### CSF proteomic data quality control

Cerebrospinal fluid (CSF) samples were collected via lumbar puncture (LP) in the morning after an overnight fast. All samples underwent the identical protocols for preparation, processing, and storage at −80°C. Proteomic data in CSF for Knight-ADRC, ADNI, FACE, Barcelona-1, and DIAN were quantified using SomaScan v4.1, while proteomic data for PPMI were quantified using SomaScan v4. SomaScan is a high-throughput platform developed by SomaLogic (Boulder, CO)^74^ that uses single-stranded DNA aptamer to quantify CSF proteins in relative fluorescent units (RFU). SomaLogic performed normalization procedures to mitigate sample^75^. The normalization involved intra-plate bias correction using hybridization control normalization, median signal and adaptive normalization by maximum likelihood. Inter-plate bias was removed by inter-plate calibration using calibrator samples.

We further performed in-house quality control on the proteomic data, as described previously^13^. SomaDataIO (v1.8.0; https://somalogic.github.io/SomaDataIO) and Biobase (v2.42.0)^76^ packages were used. Specifically, we applied the following steps: aptamers with a scale factor exceeding 0.5 and those with coefficient of variation greater than 0.15 were excluded. Datapoints for each aptamer were set to missing if their log10 transformed values outside 1.5 times the interquartile range (IQR). Aptamers and samples over 15% missing rate were removed. Duplicate samples and aptamers targeting non-human proteins were also excluded. Due to lack of scale factor and CV for SomaScan v4, we were unable to perform the first process which removes aptamers with scale factor and CV criteria. All other steps were performed as described above. In addition, we removed SomaScan v4-specific aptamers that were not present in SomaScan v4.1.

After QC, a total of 7,008 aptamers remained for analysis. Aptamer levels were log-transformed (log10) and subsequently z-score transformed to approximate a normal distribution prior to pQTL mapping. All aptamers were annotated using Universal Protein Resource (UniProt)^77^ identifiers and Entrez Gene symbols.

### Genome-wide data, quality control, and imputation

The five cohorts (Knight-ADRC, ADNI, FACE, Barcelona-1, and DIAN) were genotyped across multiple platforms: Illumina CoreExome-24 (CoreEx), Global Screening Array-24 (GSA), NeuroX2, OmniExpress-24 (OmniEx), Human660W-Quad (X660W), and Affymetrix Axiom. We performed quality control (QC) separately for each genotype array using PLINK v1.90b6.26^78^. As a part of genotype QC steps, we (i) removed variants without chromosome or position information, insertions/deletions, variants with minor allele frequency (MAF) of 0 (i.e., monomorphic variant in each array), and duplicate variants; (ii) included only autosomal chromosomes; (iii) excluded samples or variants with missingness over 0.02; (iv) excluded palindromic variants; (v) excluded variants not in Hardy-Weinberg equilibrium (HWE) (with P < 1×10^−6^); and (vi) lifted over genome coordinates from hg19 to the GRCh38/hg38 coordinates if needed. In addition, we checked the consistency between the phenotype sex of individuals and that estimated by genotype data and excluded those individuals with inconsistent sex information. We subsequently performed imputation using the TOPMed imputation server (August 2021). This manuscript used imputed variants with an imputation quality filter of Rsq > 0.30. For PPMI, we downloaded whole-genome sequencing data in VCF format (buildGRCh38/hg38). Low-quality variants and samples with a call rate of <98% were removed from our analyses. All of these QCed data were merged into one dataset.

We further performed a relatedness check using pairwise genome-wide estimates of proportion identity-by-descent. To select unrelated individuals, we retained only one sample with the higher call rate from each (either cryptically or genetically) related pair (with Pihat ≥ 0.25). Finally, we performed principal component (PC) analysis anchored by the sequencing data from the 1000 Genomes Project Phase 3^79^. We retained the 3,353 individuals of European descent. The top 10 PCs were subsequently used as covariates in the GWAS analysis to correct for any possible bias due to population stratification.

Only autosomal genetic variants were included in GRCh38/hg38 coordinates. The variants with minor allele count (MAC) > 10 were included in our analyses. In addition, we removed palindromic variants to avoid spurious associations due to unclear genotyping array strands.

### Identification of sex-specific pQTLs

To identify sex-specific or sex-biased effects of genetic variants on protein levels, we performed sex-stratified GWAS for each of 7,008 aptamers in 1,713 males and 1,640 females, separately, with the following mean model:

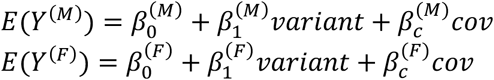

where 𝛽_%_ corresponds to the magnitude of genetic association for the aptamer. 𝑌 denotes the z-score transformed aptamer level. We included age at CSF draw, cohorts/genotype platforms, and the first 10 ancestry PCs as a set of covariates (𝑐𝑜𝑣). The analysis was conducted using PLINK2 (v2.00a2.3LM AVX2 Intel)^80^.

Significant pQTLs were identified for each sex, using two different thresholds, one for cis and another for trans signal. A cis-pQTL of a protein was defined if the index variant of a given locus was genome-wide significant (P < 5×10⁻⁸) and within 1 megabase (Mb) of the transcription start site (TSS) of the gene targeting that protein. A trans-pQTL was defined if the index variant was outside 1 Mb of TSS and was significant at P < 3.45×10⁻¹¹. This threshold corresponds to 5×10⁻⁸/1450, the Bonferroni correction for 1,450 proteomic PCs that accounted for 95% of the variance.

Once pQTLs were identified in each of the sex-stratified analyses separately, each signal was classified into five groups (Supplementary Fig 1a). If a pQTL was detected only in males, it was classified as male-specific. If it was detected only in females, it was classified as female-specific. If a pQTL was detected in both sexes, then we tested whether their effects were different. Specifically, we computed the standardized difference of genetic effects in males and females:

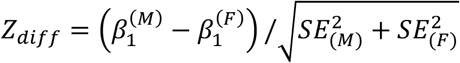

and tested whether this standardized difference (𝑍_()**_)was statistically significant at P < 0.05. If not significant, then it was classified as shared. If it was significant and if the absolute effect in males was larger, it was classified as male-biased. If it was significant and the absolute effect in females was larger, it was classified as female-biased.

All identified pQTLs were annotated using the Ensemble Variant Effect Predictor (VEP) release 107 (GRCh38.p13 assembly for Homo_sapiens)^81^ to obtain the rsID number, functional consequence (such as protein sequence alterations), and its affecting gene(s), if any.

### Replication in Stanford ADRC

To validate the identified pQTLs, we performed additional pQTL analyses in 70 males and 95 females, separately, from the Stanford Alzheimer’s Disease Research Center (Stanford ADRC). This cohort is an ongoing longitudinal study of individuals with AD and cognitively unimpaired controls. CSF proteomic data were quantified using the SomaScan v4 platform. These samples were similarly selected for European unrelated individuals. Of the index pQTLs identified in this manuscript, 1,194 were available for the matching protein and the genetic variant in the Stanford dataset. Sex-stratified genome-wide analyses were performed using PLINK v2.0, adjusting for age and the first ten ancestry principal components.

### Identification and characterization of pleiotropic regions

To identify a pleotropic region, i.e., a genomic region affecting multiple proteins, we created a locusID (shown in Supplementary Table 1) based on the distance. If each index pQTL variant resided within 1 Mb of any nearby index pQTL variants, then they were assigned to the same locusID. Within each locusID, we obtained the associated aptamers and proteins (labeled as Entrez Gene symbols; in Supplementary Table 8). They were further split into five groups (male-specific, female-specific, male-biased, female-biased, and shared).

For each of two pleotropic regions, we generated a Circos plot to visualize the association for multiple proteins using the circlize R package. The genetic variants and affecting proteins were plotted along the chromosome ideogram. Each connecting line was colored by group and weighted by the magnitude of the genetic effect size. Shared pQTLs (without any sex differences in their genetic regulations) were excluded to present those sex-dependent regulations. Gene names were displayed on the inner ring of the plot to highlight target proteins regulated within the region.

### Cell-type specificity

Cell-type specificity was performed for the proteins regulated in a sex-dependent way. Specifically, those proteins regulated by each pleotropic region were examined. We also examined cell-type specificity for the male set (including male-specific and male-biased regulations) and the female set (female-specific and biased). We downloaded the published gene expression data from human astrocytes, neurons, oligodendrocytes, microglia/macrophages, and endothelial cells^82^. We curated the expression values for all protein-coding genes targeted by aptamers in the SOMAscan 7k proteomic panel. To assess the enrichment of the observed cell-type composition of an input protein set, significance was determined using a hypergeometric test. All proteins in the SOMAscan panel were considered as a background.

### Pathway enrichment analysis

Pathway enrichment analyses for male and female sets were performed using Metascape (https://metascape.org), an integrative web-based platform. Metascape^75^ incorporates multiple biological knowledge bases, including Gene Ontology (GO) Biological Processes, GO Molecular Function, GO Cellular Components, KEGG, Reactome, and Wikipathways. The male set included 301 genes targeting proteins regulated in a male-specific and male-biased way.

Similarly, the female set included 220 genes (for female-biased and female-specific genetic regulations). The background gene set comprised all proteins assayed by the SomaScan 7K platform (n = 7,008). After removing duplicates and mapping to unique Entrez Gene Symbols, 6,161 genes were retained for use as the background set for the enrichment analysis. For each gene set, enrichment analysis was conducted using Metascape’s default pipeline, which applies a hypergeometric test followed by Benjamini–Hochberg correction for multiple hypothesis testing^75^. Enriched terms with a p-value < 1.00, a minimum count of 3 genes, and an enrichment factor > 1.5 were retained.

### Proteome-wide association study (PWAS)

We applied the FUSION framework to perform sex-stratified PWAS^21^ for sex-stratified pQTLs results. Predictive models of protein abundance were trained separately for males and females using individual-level genome-wide and proteomic data. For each analyte with significant cis-pQTLs in either sex, we extracted variants located within one Mb of the index variant. We trained three types of models, top1, lasso, and elastic net (enet), to estimate genetically regulated protein levels. Model training included the following covariates: sample IDs, age, the top 10 genetic PCs, and binary indicators for cohort/genotyping platforms. Analytes with negative heritability estimates (hsq <0) were excluded from weight computation. For each sex, we generated weight files and used them to test for association with AD and PD using GWAS summary statistics from Bellengeuz et al.^23^ for AD and Nalls et al.^24^ for PD. PWAS association testing was conducted with FUSION.assoc_test.R, using a minimum predictive R² threshold of zero to include all available models.

### Colocalization analyses

We performed Bayesian colocalization analysis^22^ to test whether a cis-pQTL signal, in males and females, separately, would share the same causal variant with each AD risk locus. Colocalization with PD was performed similarly. We examined colocalization for all cis-pQTLs significant in either sex. The coloc.abf function in the coloc R package was used with default priors (P1=1×10^-4^, P2=1×10^-4^ and P12=1×10^-5^). We considered PP.H4 > 0.5 as evidence of a shared causal variant between protein abundance and disease risk. We defined sex-specific colocalizations as those with strong evidence (PP.H4 > 0.5) in one sex, but no evidence in the other.

## Data Availability

All data supporting the findings of this study are available in this manuscript.

## Acknowledgements

We are grateful to all participants and their families, and to the contributing cohorts, institutions, and their dedicated staff.

This work was supported by grants from the National Institutes of Health, R01 AG074007 (YJS), R01 AG044546 (CC), P01 AG003991 (CC, JCM), RF1 AG053303 (CC), RF1 AG058501 (CC), U01 AG058922 (CC), P30 AG066444 (JCM), P01 AG026276 (JCM), R00 AG075238, (MEB) and the Chan Zuckerberg Initiative (CZI), the Michael J. Fox Foundation (CC), the Alzheimer’s Association Zenith Fellows Award (ZEN-22-848604, awarded to CC).

<u>ADNI acknowledgement:</u> Data collection and sharing for this project were funded by the Alzheimer’s Disease Neuroimaging Initiative (ADNI) (National Institutes of Health Grant U01 AG024904) and DOD ADNI (Department of Defense award number W81XWH-12-2-0012). ADNI is funded by the National Institute on Aging, the National Institute of Biomedical Imaging and Bioengineering, and through generous contributions from the following: AbbVie, Alzheimer’s Association; Alzheimer’s Drug Discovery Foundation; Araclon Biotech; BioClinica, Inc.; Biogen; Bristol-Myers Squibb Company; CereSpir, Inc.; Cogstate; Eisai Inc.; Elan Pharmaceuticals, Inc.; Eli Lilly and Company; EuroImmun; F. Hoffmann-La Roche Ltd and its affiliated company Genentech, Inc.; Fujirebio; GE Healthcare; IXICO Ltd.; Janssen Alzheimer Immunotherapy Research & Development, LLC.; Johnson & Johnson Pharmaceutical Research & Development LLC.; Lumosity; Lundbeck; Merck & Co., Inc.; Meso Scale Diagnostics, LLC.; NeuroRx Research; Neurotrack Technologies; Novartis Pharmaceuticals Corporation; Pfizer Inc.; Piramal Imaging; Servier; Takeda Pharmaceutical Company; and Transition Therapeutics. The Canadian Institutes of Health Research is providing funds to support ADNI clinical sites in Canada. Private sector contributions are facilitated by the Foundation for the National Institutes of Health (www.fnih.org). The grantee organization is the Northern California Institute for Research and Education, and the study is coordinated by the Alzheimer’s Therapeutic Research Institute at the University of Southern California. ADNI data are disseminated by the Laboratory for Neuro Imaging at the University of Southern California.

## References

1. Rajan, K.B. et al. Population estimate of people with clinical Alzheimer’s disease and mild cognitive impairment in the United States (2020-2060). Alzheimers Dement 17, 1966–1975 (2021).

2. Del-Aguila, J.L. et al. Assessment of the Genetic Architecture of Alzheimer’s Disease Risk in Rate of Memory Decline. J Alzheimers Dis 62, 745–756 (2018).

3. Cattaneo, C. & Pagonabarraga, J. Sex Differences in Parkinson’s Disease: A Narrative Review. Neurol Ther 14, 57–70 (2025).

4. Lopez-Lee, C., Torres, E.R.S., Carling, G. & Gan, L. Mechanisms of sex differences in Alzheimer’s disease. Neuron 112, 1208–1221 (2024).

5. Tranchevent, L.C., Halder, R. & Glaab, E. Systems level analysis of sex-dependent gene expression changes in Parkinson’s disease. NPJ Parkinsons Dis 9, 8 (2023).

6. Deming, Y. et al. Sex-specific genetic predictors of Alzheimer’s disease biomarkers. Acta Neuropathol 136, 857–872 (2018).

7. Aberg, D. et al. Increased Cerebrospinal Fluid Level of Insulin-like Growth Factor-II in Male Patients with Alzheimer’s Disease. J Alzheimers Dis 48, 637–46 (2015).

8. Piccio, L. et al. Cerebrospinal fluid soluble TREM2 is higher in Alzheimer disease and associated with mutation status. Acta Neuropathol 131, 925–33 (2016).

9. Viswanathan, J. et al. An association study between granulin gene polymorphisms and Alzheimer’s disease in Finnish population. Am J Med Genet B Neuropsychiatr Genet 150B, 747–50 (2009).

10. Oliva, M. et al. The impact of sex on gene expression across human tissues. Science 369(2020).

11. Wingo, A.P. et al. Sex differences in brain protein expression and disease. Nat Med 29, 2224–2232 (2023).

12. Koprulu, M. et al. Sex differences in the genetic regulation of the human plasma proteome. Nat Commun 16, 4001 (2025).

13. Western, D. et al. Proteogenomic analysis of human cerebrospinal fluid identifies neurologically relevant regulation and implicates causal proteins for Alzheimer’s disease. Nat Genet 56, 2672–2684 (2024).

14. Yang, C. et al. Genomic atlas of the proteome from brain, CSF and plasma prioritizes proteins implicated in neurological disorders. Nat Neurosci 24, 1302–1312 (2021).

15. Wolosowicz, M., Prokopiuk, S. & Kaminski, T.W. The Complex Role of Matrix Metalloproteinase-2 (MMP-2) in Health and Disease. Int J Mol Sci 25(2024).

16. Kanda, H., Shimamura, R., Koizumi-Kitajima, M. & Okano, H. Degradation of Extracellular Matrix by Matrix Metalloproteinase 2 Is Essential for the Establishment of the Blood-Brain Barrier in Drosophila. iScience 16, 218–229 (2019).

17. Nikolakopoulou, A.M., Dutta, R., Chen, Z., Miller, R.H. & Trapp, B.D. Activated microglia enhance neurogenesis via trypsinogen secretion. Proc Natl Acad Sci U S A 110, 8714–9 (2013).

18. Schondorf, D.C. et al. iPSC-derived neurons from GBA1-associated Parkinson’s disease patients show autophagic defects and impaired calcium homeostasis. Nat Commun 5, 4028 (2014).

19. Medoh, U.N. et al. The Batten disease gene product CLN5 is the lysosomal bis(monoacylglycero)phosphate synthase. Science 381, 1182–1189 (2023).

20. Henderson, A.D. & Miller, N.R. Carotid-cavernous fistula: current concepts in aetiology, investigation, and management. Eye (Lond) 32, 164–172 (2018).

21. Gusev, A. et al. Integrative approaches for large-scale transcriptome-wide association studies. Nat Genet 48, 245–52 (2016).

22. Giambartolomei, C. et al. Bayesian test for colocalisation between pairs of genetic association studies using summary statistics. PLoS Genet 10, e1004383 (2014).

23. Bellenguez, C. et al. New insights into the genetic etiology of Alzheimer’s disease and related dementias. Nat Genet 54, 412–436 (2022).

24. Blauwendraat, C. et al. Investigation of Autosomal Genetic Sex Differences in Parkinson’s Disease. Ann Neurol 90, 35–42 (2021).

25. Paranjpe, M.D. et al. Sex-Specific Cross Tissue Meta-Analysis Identifies Immune Dysregulation in Women With Alzheimer’s Disease. Front Aging Neurosci 13, 735611 (2021).

26. Luo, H. et al. Gene Expression Profiling Reveals Potential Players of Sex Determination and Asymmetrical Development in Chicken Embryo Gonads. Int J Mol Sci 24(2023).

27. Ratnu, V.S., Emami, M.R. & Bredy, T.W. Genetic and epigenetic factors underlying sex differences in the regulation of gene expression in the brain. J Neurosci Res 95, 301–310 (2017).

28. Bridges, J., Ramirez-Guerrero, J.A. & Rosa-Garrido, M. Gender-specific genetic and epigenetic signatures in cardiovascular disease. Front Cardiovasc Med 11, 1355980 (2024).

29. Pietzner, M. et al. Mapping the proteo-genomic convergence of human diseases. Science 374, eabj1541 (2021).

30. Dessy, T. et al. Disentangling the effects of sex and gender on APOE varepsilon4-related neurocognitive impairment. Alzheimers Dement (Amst) 17, e70111 (2025).

31. Hohman, T.J. et al. Sex-Specific Association of Apolipoprotein E With Cerebrospinal Fluid Levels of Tau. JAMA Neurol 75, 989–998 (2018).

32. Xu, X. et al. Sex Differences in Apolipoprotein E and Alzheimer Disease Pathology Across Ancestries. JAMA Netw Open 8, e250562 (2025).

33. Zhang, Z.D. et al. RNF115 Inhibits the Post-ER Trafficking of TLRs and TLRs-Mediated Immune Responses by Catalyzing K11-Linked Ubiquitination of RAB1A and RAB13. Adv Sci (Weinh) 9, e2105391 (2022).

34. Zhou, Z. et al. Integrative genomic analysis of PPP3R1 in Alzheimer’s disease: a potential biomarker for predictive, preventive, and personalized medical approach. EPMA J 12, 647–658 (2021).

35. Wang, H. et al. Neurofilament proteins in axonal regeneration and neurodegenerative diseases. Neural Regen Res 7, 620–6 (2012).

36. Mendsaikhan, A., Tooyama, I. & Walker, D.G. Microglial Progranulin: Involvement in Alzheimer’s Disease and Neurodegenerative Diseases. Cells 8(2019).

37. Nalls, M.A. et al. Evidence for GRN connecting multiple neurodegenerative diseases. Brain Commun 3, fcab095 (2021).

38. Akiyama, T., Raftery, L.A. & Wharton, K.A. Bone morphogenetic protein signaling: the pathway and its regulation. Genetics 226(2024).

39. Van Aerde, N., Van den Berghe, G. & Hermans, G. Weakness in the ICU: the right weight on the right scale. Intensive Care Med 47, 137–138 (2021).

40. Shah, T.A. & Rogers, M.B. Unanswered Questions Regarding Sex and BMP/TGF-beta Signaling. J Dev Biol 6(2018).

41. Porro, C., Cianciulli, A. & Panaro, M.A. The Regulatory Role of IL-10 in Neurodegenerative Diseases. Biomolecules 10(2020).

42. Allavena, R., Noy, S., Andrews, M. & Pullen, N. CNS elevation of vascular and not mucosal addressin cell adhesion molecules in patients with multiple sclerosis. Am J Pathol 176, 556–62 (2010).

43. Pyka-Fosciak, G., Lis, G.J. & Litwin, J.A. Adhesion Molecule Profile and the Effect of Anti-VLA-4 mAb Treatment in Experimental Autoimmune Encephalomyelitis, a Mouse Model of Multiple Sclerosis. Int J Mol Sci 23(2022).

44. Wang, Z. et al. QSP Modeling Shows Pathological Synergism Between Insulin Resistance and Amyloid-Beta Exposure in Upregulating VCAM1 Expression at the BBB Endothelium. CPT Pharmacometrics Syst Pharmacol 14, 561–571 (2025).

45. Tedde, A. et al. Interleukin-10 promoter polymorphisms influence susceptibility to ulcerative colitis in a gender-specific manner. Scand J Gastroenterol 43, 712–8 (2008).

46. Stagi, M., Klein, Z.A., Gould, T.J., Bewersdorf, J. & Strittmatter, S.M. Lysosome size, motility and stress response regulated by fronto-temporal dementia modifier TMEM106B. Mol Cell Neurosci 61, 226–40 (2014).

47. Perneel, J. et al. Accumulation of TMEM106B C-terminal fragments in neurodegenerative disease and aging. Acta Neuropathologica 145, 285–302 (2022).

48. Feng, T., et al. Loss of TMEM106B and PGRN leads to severe lysosomal abnormalities and neurodegeneration in mice. EMBO Reports 21(2020).

49. Lüningschrör, P. et al. The FTLD Risk Factor TMEM106B Regulates the Transport of Lysosomes at the Axon Initial Segment of Motoneurons. Cell reports 30 10, 3506–3519 (2020).

50. Feng, T., Lacrampe, A. & Hu, F. Physiological and pathological functions of TMEM106B: a gene associated with brain aging and multiple brain disorders. Acta Neuropathologica, 1–13 (2021).

51. Jiao, H.-S., Yuan, P. & Yu, J.-T. TMEM106B aggregation in neurodegenerative diseases: linking genetics to function. Molecular Neurodegeneration 18, 54 (2023).

52. Hu, Y. et al. rs1990622 variant associates with Alzheimer’s disease and regulates TMEM106B expression in human brain tissues. BMC Medicine 19(2021).

53. Yan, H. et al. Assessing the impact of novel risk loci on Alzheimer’s and Parkinson’s diseases in a Chinese Han cohort. Frontiers in Neurology 15(2024).

54. Wang, L.Y. et al. Associations among Angiotensin-Converting Enzyme, Neuroinflammation, and Cerebrospinal Fluid Biomarkers of Alzheimer’s Disease in Non-Dementia Adults. Neurotox Res 43, 20 (2025).

55. Santiago, T.C. et al. Angiotensin-converting enzymes as druggable features of psychiatric and neurodegenerative disorders. Journal of Neurochemistry 166, 138–155 (2023).

56. Hu, J., Igarashi, A., Kamata, M. & Nakagawa, H. Angiotensin-converting Enzyme Degrades Alzheimer Amyloid β-Peptide (Aβ); Retards Aβ Aggregation, Deposition, Fibril Formation; and Inhibits Cytotoxicity*. Journal of Biological Chemistry 276, 47863–47868 (2001).

57. He, M., Zhang, F., Qi, J. & Zhang, W. Missense mutation of angiotensin converting enzyme gene in an Alzheimer’s disease patient: a case report. Front Neurosci 18, 1343279 (2024).

58. Ni, J., Xiao, S., Li, X. & Sun, L. ACE gene missense mutation in a case with early-onset, rapid progressing dementia. Gen Psychiatr 32, e100028 (2019).

59. Bonneh-Barkay, D. & Wiley, C.A. Brain Extracellular Matrix in Neurodegeneration. Brain Pathology 19, 573–585 (2009).

60. Moretto, E., Stuart, S., Surana, S., Vargas, J.N.S. & Schiavo, G. The Role of Extracellular Matrix Components in the Spreading of Pathological Protein Aggregates. Frontiers in Cellular Neuroscience **Volume** 16 **-**2022(2022).

61. Freese, C. et al. A Novel Blood-Brain Barrier Co-Culture System for Drug Targeting of Alzheimer’s Disease: Establishment by Using Acitretin as a Model Drug. PLOS ONE 9, e91003 (2014).

62. de Rus Jacquet, A., et al. The contribution of inflammatory astrocytes to BBB impairments in a brain-chip model of Parkinson’s disease. Nature Communications 14, 3651 (2023).

63. Riboldi, G.M. & Di Fonzo, A.B. GBA, Gaucher Disease, and Parkinson’s Disease: From Genetic to Clinic to New Therapeutic Approaches. Cells 8(2019).

64. O’Regan, G., deSouza, R.M., Balestrino, R. & Schapira, A.H. Glucocerebrosidase Mutations in Parkinson Disease. J Parkinsons Dis 7, 411–422 (2017).

65. Milenkovic, I., Blumenreich, S. & Futerman, A.H. GBA mutations, glucosylceramide and Parkinson’s disease. Curr Opin Neurobiol 72, 148–154 (2022).

66. Smith, L. & Schapira, A.H.V. GBA Variants and Parkinson Disease: Mechanisms and Treatments. Cells 11(2022).

67. Ortega, R.A. et al. Differences in Sex-Specific Frequency of Glucocerebrosidase Variant Carriers and Familial Parkinsonism. Mov Disord 37, 2217–2225 (2022).

68. Kukurba, K.R. et al. Impact of the X Chromosome and sex on regulatory variation. Genome Res 26, 768–77 (2016).

69. Schurz, H. et al. The X chromosome and sex-specific effects in infectious disease susceptibility. Human Genomics 13, 2 (2019).

70. Fernandez, M.V. et al. Genetic and multi-omic resources for Alzheimer disease and related dementia from the Knight Alzheimer Disease Research Center. Sci Data 11, 768 (2024).

71. Mueller, S.G. et al. Ways toward an early diagnosis in Alzheimer’s disease: the Alzheimer’s Disease Neuroimaging Initiative (ADNI). Alzheimers Dement 1, 55–66 (2005).

72. Moreno-Grau, S. et al. Genome-wide association analysis of dementia and its clinical endophenotypes reveal novel loci associated with Alzheimer’s disease and three causality networks: The GR@ACE project. Alzheimers Dement 15, 1333–1347 (2019).

73. Marek, K. et al. The Parkinson’s progression markers initiative (PPMI) - establishing a PD biomarker cohort. Ann Clin Transl Neurol 5, 1460–1477 (2018).

74. Gold, L. et al. Aptamer-based multiplexed proteomic technology for biomarker discovery. PLoS One 5, e15004 (2010).

75. Zhou, Y. et al. Metascape provides a biologist-oriented resource for the analysis of systems-level datasets. Nat Commun 10, 1523 (2019).

76. Huber, W. et al. Orchestrating high-throughput genomic analysis with Bioconductor. Nat Methods 12, 115–21 (2015).

77. UniProt, C. UniProt: the universal protein knowledgebase in 2021. Nucleic Acids Res 49, D480–D489 (2021).

78. Purcell, S. et al. PLINK: a tool set for whole-genome association and population-based linkage analyses. Am J Hum Genet 81, 559–75 (2007).

79. Genomes Project, C., et al. A global reference for human genetic variation. Nature 526, 68–74 (2015).

80. Chang, C.C. et al. Second-generation PLINK: rising to the challenge of larger and richer datasets. Gigascience 4, 7 (2015).

81. McLaren, W. et al. The Ensembl Variant Effect Predictor. Genome Biol 17, 122 (2016).

82. Zhang, Y. et al. Purification and Characterization of Progenitor and Mature Human Astrocytes Reveals Transcriptional and Functional Differences with Mouse. Neuron 89, 37–53 (2016).

